# Immunogenicity of heterologous BNT162b2 booster in fully vaccinated individuals with CoronaVac against SARS-CoV-2 variants Delta and Omicron: the Dominican Republic Experience

**DOI:** 10.1101/2021.12.27.21268459

**Authors:** Eddy Pérez-Then, Carolina Lucas, Valter Silva Monteiro, Marija Miric, Vivian Brache, Leila Cochon, Chantal B. F. Vogels, Elena De la Cruz, Aidelis Jorge, Margarita De los Santos, Patricia Leon, Mallery I. Breban, Kendall Billig, Inci Yildirim, Claire Pearson, Randy Downing, Emily Gagnon, Anthony Muyombwe, Jafar Razeq, Melissa Campbell, Albert Ko, Saad B. Omer, Nathan D. Grubaugh, Sten H. Vermund, Akiko Iwasaki

## Abstract

The recent emergence of the SARS-CoV-2 Omicron variant is raising concerns because of its increased transmissibility and by its numerous spike mutations with potential to evade neutralizing antibodies elicited by COVID-19 vaccines. The Dominican Republic was among the first countries in recommending the administration of a third dose COVID-19 vaccine to address potential waning immunity and reduced effectiveness against variants. Here, we evaluated the effects of a heterologous BNT162b2 mRNA vaccine booster on the humoral immunity of participants that had received a two-dose regimen of CoronaVac, an inactivated vaccine used globally. We found that heterologous CoronaVac prime followed by BNT162b2 booster regimen induces elevated virus-specific antibody levels and potent neutralization activity against the ancestral virus and Delta variant, resembling the titers obtained after two doses of mRNA vaccines. While neutralization of Omicron was undetectable in participants that had received a two-dose regimen of CoronaVac vaccine, BNT162b2 booster resulted in a 1.4-fold increase in neutralization activity against Omicron, compared to two-dose mRNA vaccine. Despite this increase, neutralizing antibody titers were reduced by 6.3-fold and 2.7-fold for Omicron compared to ancestral and Delta variant, respectively. Surprisingly, previous SARS-CoV-2 infection did not affect the neutralizing titers for Omicron in participants that received the heterologous regimen. Our findings have immediate implications for multiples countries that previously used a two-dose regimen of CoronaVac and reinforce the notion that the Omicron variant is associated with immune escape from vaccines or infection-induced immunity, highlighting the global need for vaccine boosters to combat the impact of emerging variants.

## Introduction

The ongoing evolution of SARS-CoV-2 and recent emergence of Omicron variant raise concerns about its increased transmissibility and vaccine effectiveness. CoronaVac COVID-19 vaccine is a 2-dose β-propiolactone-inactivated, aluminium hydroxide-adjuvanted COVID-19 vaccine administered on a 0/14-28-day schedule. CoronaVac is widely used globally and has been authorized in 48 countries, with 85% and 80% of effectiveness against hospital admission and death, respectively (World Health Organization, https://www.who.int/). However, with the emergence of new SARS-CoV-2 variants and the waning immunity of vaccines over time, multiple countries initiated the administration of booster doses (Goldberg et al., 2021; Naaber et al., 2021; Pegu et al., 2021). The Dominican Republic was among the first countries in recommending the administration of a third dose COVID-19 vaccine as a booster dose to address potential waning immunity and reduced effectiveness against variants.

The Omicron variant contains up to 36 mutations in spike protein (Pulliam et al., 2021) rendering many vaccines less effective. Omicron variant is also highly transmissible, overtaking Delta as the dominant variant in many countries. To assess the potential risk of vaccine immune evasion we assembled a cohort of CoronaVac-vaccinated individuals that received a heterologous BNT162b2 mRNA vaccine boost. We investigated vaccine-induced neutralizing antibody titers against the Delta (B.1.617.2 lineage) and Omicron (BA.1 lineage) variants and compared them to the ancestral A lineage, using authentic SARS-CoV-2 isolates.

### Heterologous vaccination induces elevated antibody titers

To characterize SARS-CoV-2-specific adaptive immune responses, we analyzed plasma samples from 101 participants who received the BNT162b2 booster dose at least four weeks after the second dose of CoronaVac vaccine between July 30 and August 27, 2021. Plasma samples were collected longitudinally at Departamento de Investigaciones Biomedicas, Clinica Evangelina Rodriguez, PROFAMILIA, Santo Domingo, Dominican Republic, at baseline (prior to booster), 7 and 28 days after de booster (third dose) administration and were subjected to ELISA and neutralization assays using authentic virus. Data from a previous cohort, composed of healthcare workers (HCWs) from the Yale-New Haven Hospital (YNHH) that received two doses of mRNA COVID-19 vaccines (mRNA-1273, Moderna or BNT162b2, Pfizer-BioNTech) were used as reference (Lucas et al., 2021). The mean ages of the participants (majority females, 70%) was 40.4 ± 13.4 and their body mass index was 27.5 ± 5.5. Cohort demographics, vaccination status and serostatus are summarized in Table 1.

**Table 1.** Patient’s demographic data Dominican Republic.

**1.1. Dominican Republic Cohort**
| Individual variables | N | (%) | Mean | SD |
| --- | --- | --- | --- | --- |
| Age | 101 |  | 40.4 | 13.4 |
| Gender |  |  |  |  |
| Male | 31 | 30.7 |  |  |
| Female | 70 | 69.3 |  |  |
| BMI | 101 |  | 27.63 | 5.53 |
| Infection Status |  |  |  |  |
| Non-previous infected | 75 | 74.25 |  |  |
| Previous infected | 26 | 25.74 |  |  |
| Days since last infection |  |  | 326.26 | 131.01 |
| Days since last SinoVac dose | 101 |  | 113 | 33 |

**1.2. HCWs-Yale Cohort**
| Individual variables | N | (%) | Mean | SD |
| --- | --- | --- | --- | --- |
| Age | 37 |  | 44.9 | 12.5 |
| Gender |  |  |  |  |
| Male | 7 | 18.9 |  |  |
| Female | 30 | 81.1 |  |  |
| Infection Status |  |  |  |  |
| Non-previous infected | 18 | 48.65 |  |  |
| Previous infected | 19 | 51.35 |  |  |
| Vaccine | 101 |  | 113 | 33 |
| BNT162b2 | 7 | 18.91 |  |  |
| mRNA-1273 | 30 | 81.09 |  |  |

Plasma antibody reactivity to Spike protein and receptor binding domain (RBD) of SARS-CoV-2 were measured at baseline, 7 and 28 days post BNT162b2 booster. Virus-specific IgG titers increased at 7 days over baseline, and were further elevated on day 28 post booster shot (Figure 1a,b). CoronaVac fully vaccinated individuals who received the BNT162b2 booster developed high anti-RBD IgG titers that reached equivalent levels as the HCW who received 2 doses of BNT162b2, 28 days earlier (Lucas et al., 2021).

**Figure 1:**
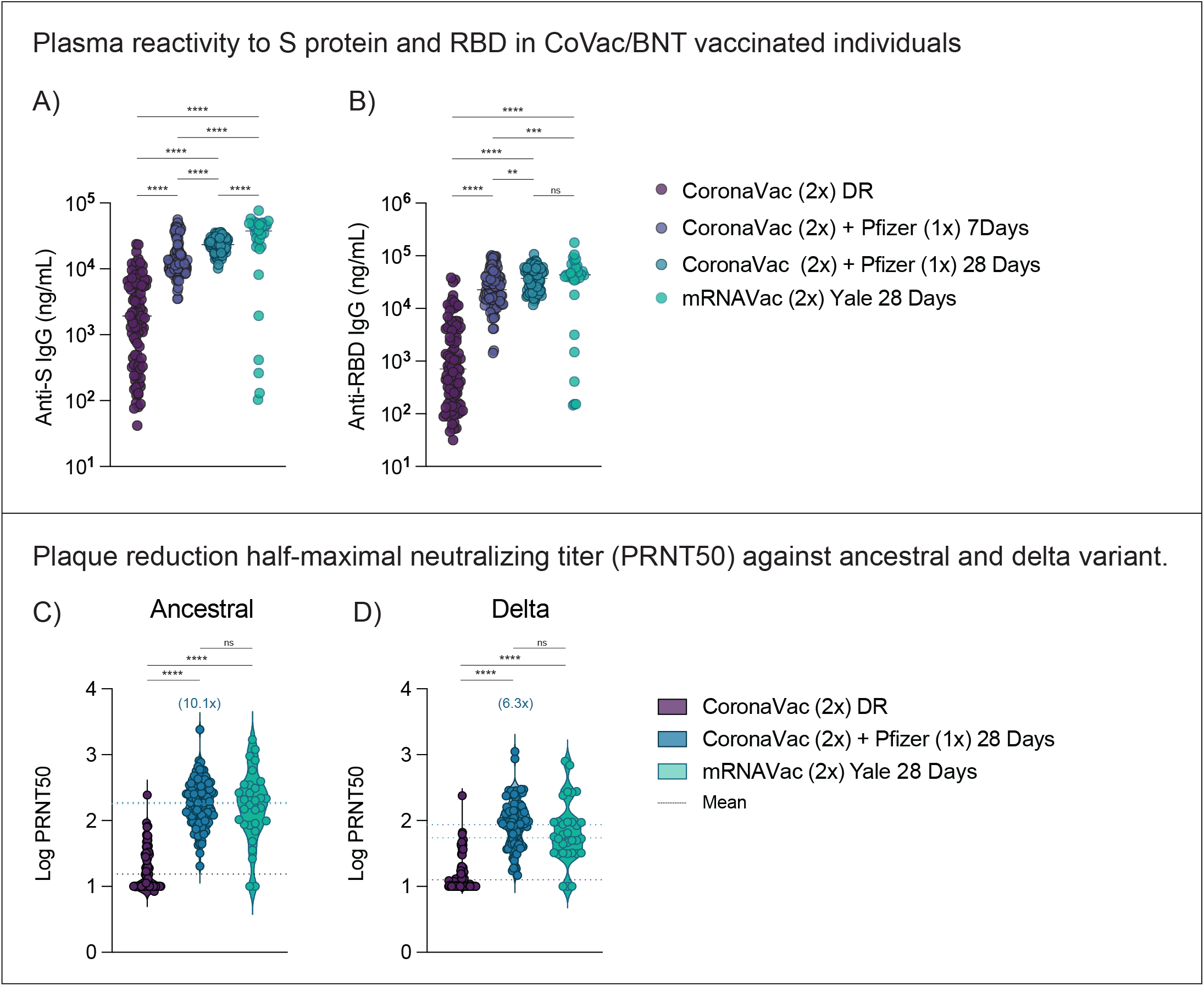
Characterization of vaccine induced immunity post heterologous CoronaVac/ BNT162b2 vaccination. Dominican Republic participants received 2 doses of CoronaVac vaccine followed by a heterologous booster with BNT162b2 mRNA vaccine. Plasma samples were collected as indicated: Baseline, prior to vaccination booster (CoronaVac (2x) DR), -7 and -28 days post 3 dose (CoronaVac (2x) + Pfizer (1x). HCW participants received 2 doses of the mRNA vaccine and plasma samples were used for comparison (mRNAVac (2x) Yale) **a, b**, Plasma reactivity to S protein and RBD measured over time by ELISA. Anti-S (a) and Anti-RBD (b) IgG levels comparison in vaccinated participants after heterologous booster. S, spike. RBD, receptor binding domain. Significance was assessed by One-way ANOVA corrected for multiple comparisons using Tukey’s method. Horizontal lines indicate median values. **c, d**, Neutralization assay using wild-type SARS-CoV-2. Plasma neutralization capacity against ancestral strain (WA1, USA) (**c**) and B.1.617.2 (Delta variant) (**d)** in vaccinated participants at the baseline (prior to booster, purple) or 28 days post 3 dose (blue). Significance was assessed by One-way ANOVA corrected for multiple comparisons using Tukey’s method. Violin plots represent mean values ± standard deviations. Horizontal lines indicate mean values and were coloured accordingly. The numbers in parentheses indicate the median fold change in neutralization resistance for the indicated variants for participants post booster vaccination. n=101 in each respective time point. Each dot represents a single individual. ****p < .0001 ***p < .001 **p < .01*p < .05.

### Heterologous vaccination induces neutralizing antibodies against VOCs

We then measured the ability of serum samples to neutralize SARS-CoV-2, lineage A (ancestral strain, USA-WA1/2020) and B.1.617.2 (Delta variant). Individuals who were fully vaccinated with CoronaVac and received BNT162b2 booster displayed a 10.1- and 6.3-fold increase in neutralization activity against ancestral and delta variant, respectively, 28 days after the booster shot; no statistical differences were observed in neutralization titers between the CoronaVac/BNT162b2 versus two-dose mRNA vaccinated individuals, 28 days after the last shot (Figure 1c, d).

Next, due to the recent emergence of SARS-CoV-2 Omicron, first identified in in Botswana and South Africa, we extended our analysis to investigate potential neutralizing antibody (Nab) escape post vaccination. Within our HCWs cohort composed of 2x mRNA vaccinated individuals, we observed a significantly reduced plaque reduction half-maximal neutralizing titer (PRNT50), 14.5-fold reduction against the Omicron variant (Figure 2a), at 28 days post second vaccination dose, at the peak of neutralization titers (Lucas et al, 2021).

**Figure 2:**
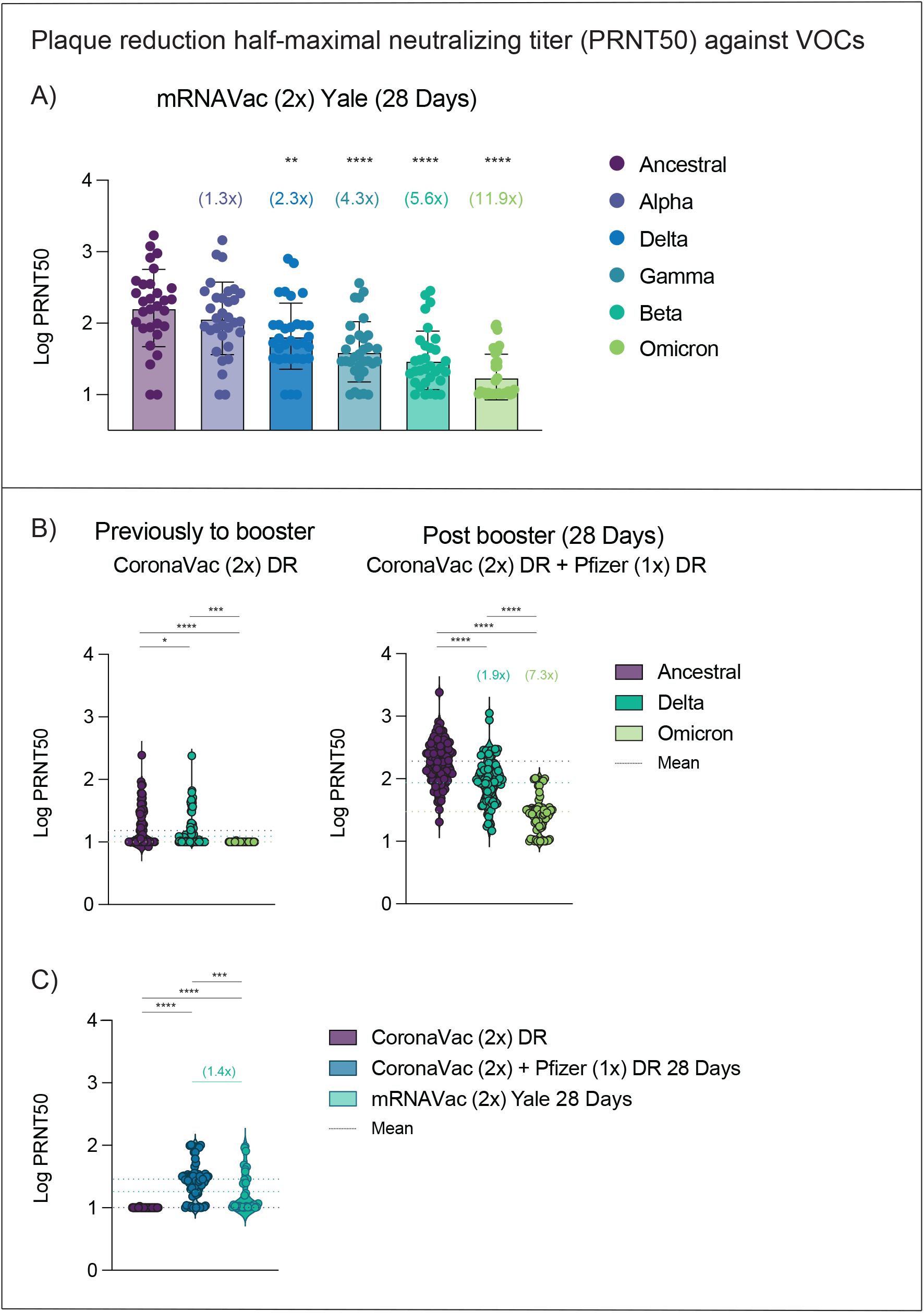
Impact of SARS-CoV-2 Omicron on neutralization capacity of CoronaVac/ BNT162b2-vaccinated participants. **a**, Plasma neutralization titers against ancestral lineage A virus, (WA1, USA) and VOCs: Alpha, Beta, Delta and Omicron. SARS-CoV-2 variants were isolated from nasopharyngeal swabs of infected individuals and ancestral (WA1, USA) isolate was obtained from BEI. Neutralization capacity was accessed using plasma samples from HCW participants that received 2 doses of the mRNA vaccine (mRNAVac (2x) Yale), 28 days post second vaccination dose at the experimental sixfold serial dilutions (from 1:3 to 1:2430). Significance was assessed by One-way ANOVA corrected for multiple comparisons using Dunnett’s method. Neutralization capacity to the variants was compared to neutralization capacity against the ancestral strain. Boxes represent mean values ± standard deviations. The numbers in parentheses indicate the median fold change in neutralization resistance for the indicated variants compared to ancestral strain. n=32/group Each dot represents a single individual. ****p < .0001 **p < .005. **b, c** Plasma neutralization titers from Dominican Republic participants that received 2 doses of CoronaVac vaccine followed by a heterologous booster with BNT162b2 mRNA vaccine. **b**, Neutralization titers against ancestral lineage A virus, (WA1, USA) and Delta and Omicron Baseline, prior to vaccination booster (left panel, CoronaVac (2x) DR), and -28 days post 3 dose (right panel, CoronaVac (2x) + Pfizer (1x). Significance was assessed by One-way ANOVA corrected for multiple comparisons using Tukey’s method. Violin plots represent mean values ± standard deviations. Horizontal lines indicate mean values and were coloured accordingly. The numbers in parentheses indicate the median fold change in neutralization resistance for the indicated variants compared to ancestral strain. n=101 (Ancestral); n=101 (Delta); n=80 (Omicron). Each dot represents a single individual. ****p < .0001 ***p < .001 *p < .05. **c**, Plasma neutralization capacity against Omicron in vaccinated participants at the baseline (prior to booster, purple) or 28 days post 3 dose (blue). HCW participants received 2 doses of the mRNA vaccine and plasma samples were used for comparison (mRNAVac (2x) Yale, green). Significance was assessed by One-way ANOVA corrected for multiple comparisons using Tukey’s method. Violin plots represent mean values ± standard deviations. Horizontal lines indicate mean values and were coloured accordingly. The numbers in parentheses indicate the median fold change in neutralization resistance between 28 days vaccinated particpants that received CoronaVac (2x) + Pfizer (1x) DR or mRNAVac (2x) Yale. n=64 CoronaVac (2x); n=80 CoronaVac (2x) + Pfizer (1x) DR; n=32 mRNAVac (2x) Yale. Each dot represents a single individual. ****p < .0001 ***p < .001.

Recent studies demonstrate that booster doses of homologous mRNA vaccines can enhance NAb response against the Omicron variant (Basile et al., 2021; Doria-Rose et al., 2021; Garcia-Beltran et al., 2021; Gruell et al., 2021). To further access neutralization activity post heterologous CoronaVac/BNT162b2 booster regimen, we compared neutralizations titers previously and post booster vaccination against the ancestral lineage, Delta and Omicron variant. Omicron was 6.3-fold less sensitive to neutralization than ancestral and 2.7-fold less sensitive than Delta variant when assayed with plasma samples obtained 28 days post BNT162b2 booster (Figure 2b). Notably, plasma from those who received 2 doses of CoronaVac had no neutralizing antibodies to Omicron prior to booster (Figure 2b). However, after the mRNA booster, plasma from Coronavac/BNT162b2 recipients developed higher neutralizing antibody titers against the Omicron than 2x mRNA vaccinated individuals, 28 days after the last shot (Figure 2c). Despite this increase, PRNT50 values and neutralizing antibody titers are reduced (IC50 mean, 1.4; and 6.3-fold reduction) for Omicron compared to ancestral and Delta SARS-CoV-2, respectively.

### Impact of previous infection on NAb

Finally, we separated individuals by their SARS-CoV-2 previous infection status (i.e. previously infected vs uninfected) and determined their neutralization titers against Omicron post vaccination. As previously observed for other SARS-CoV-2 isolates (Lucas et al., 2021), the neutralizing antibody titers against the Omicron for previously infected individuals were higher when compared to non-previously infected individuals that received 2x mRNA vaccines, 28 days after the last shot (Figure 3a). Omicron decreased neutralization titers by 17.3 fold (compared to lineage A) in uninfected and by 10.7 fold in previously infected mRNA 2x vaccinated individuals (Figure 3a). Surprisingly, the neutralization titers are reduced for Delta variant in non-previously infected individuals but not for Omicron, in individuals that received the heterologous CoronaVac/BNT162b2 booster regimen (Figure 3b,c).

**Figure 3:**
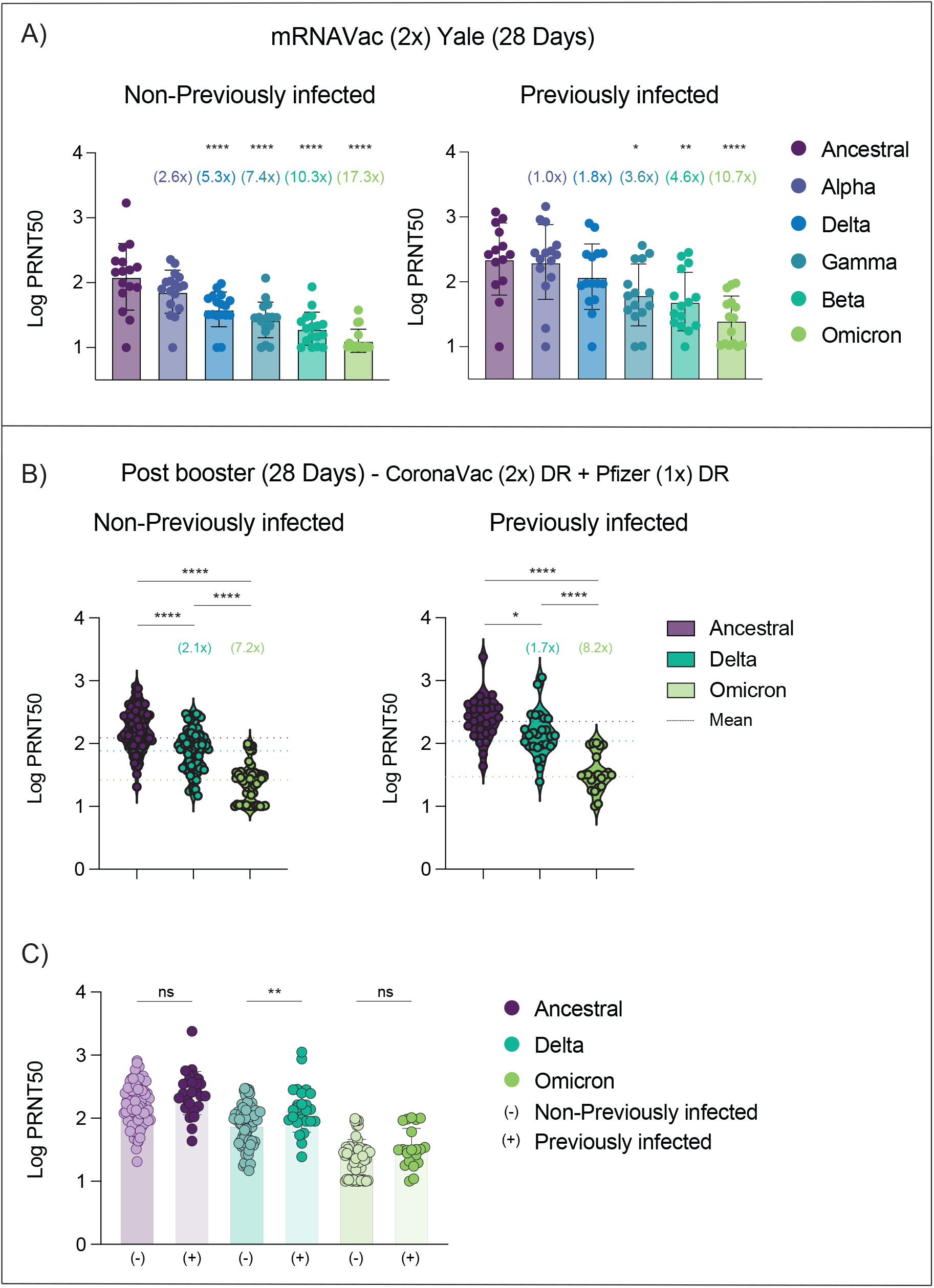
Neutralizing activity comparison in CoronaVac/BNT162b2-vaccinated participants previously infected or not to SARS-CoV-2. **a-c**, Neutralization titer comparison among vaccinated participants previously infected or not to SARS-CoV-2. **a**, Plasma neutralization titers against ancestral lineage A virus, (WA1, USA) and VOCs: Alpha, Beta, Delta and Omicron. SARS-CoV-2 variants were isolated from nasopharyngeal swabs of infected individuals and ancestral (WA1, USA) isolate was obtained from BEI. Neutralization capacity was accessed using plasma samples from HCW participants that received 2 doses of the mRNA vaccine (mRNAVac (2x) Yale), 28 days post second vaccination dose at the experimental sixfold serial dilutions (from 1:3 to 1:2430). Significance was assessed by One-way ANOVA corrected for multiple comparisons using Dunnett’s method. Neutralization capacity to the variants was compared to neutralization capacity against the ancestral strain. Boxes represent mean values ± standard deviations. The numbers in parentheses indicate the median fold change in neutralization resistance for the indicated variants compared to ancestral strain. n=32/group Each dot represents a single individual. ****p < .0001 **p < .005. *p < .05 **b, c** Plasma neutralization titers from Dominican Republic participants that received 2 doses of CoronaVac vaccine followed by a heterologous booster with BNT162b2 mRNA vaccine. **b**, Neutralization titers against ancestral lineage A virus, (WA1, USA) and Delta and Omicron -28 days post 3 dose. (left panel, non-previously infected; right panel, SARS-CoV-2 previously infected participants). Significance was assessed by One-way ANOVA corrected for multiple comparisons using Tukey’s method. Violin plots represent mean values ± standard deviations. Horizontal lines indicate mean values and were coloured accordingly. The numbers in parentheses indicate the median fold change in neutralization resistance for the indicated variants compared to ancestral strain. (Non-previously infected; n=75 (Ancestral); n=75 (Delta); n=57 (Omicron); (Previously infected; n=26 (Ancestral); n=26 (Delta); n=24 (Omicron). Each dot represents a single individual. ****p < .0001 *p < .05. **c**, Neutralization titers comparison among vaccinated participants previously infected or not to SARS-CoV-2. Significance was accessed using unpaired two-tailed t-test. Boxes represent mean values ± standard deviations. (-) Vaccinated-uninfected, n=57; (+) Vaccinated-Previously infected, n=24. Each dot represents a single individual **p < .01.

## Discussion

In vaccinated volunteers in the Dominican Republic, we found that a regimen of heterologous two-dose CoronaVac prime followed by a single BNT162b2 booster induces elevated virus-specific antibody levels and potent neutralization activity against ancestral and Delta SARS-CoV-2 strains, resembling the titers obtained after two doses of mRNA vaccines. However, the significantly reduced neutralization titers against Omicron suggest a greater risk of vaccine breakthrough infections. Clinical follow-up is needed to assess risk of serious disease in these individuals.

The Omicron variant carries multiple spike mutations that poses a potential threat to monoclonal antibody escape and vaccine effectiveness. In agreement with previous reports, our data show that Omicron variant escapes neutralizing antibodies elicited by 2x mRNA or 2x CoronaVac vaccines. Notably, none of the 2x CoronaVac recipients had detectable neutralizing antibody against SARS-CoV-2 Omicron variant. Preliminary studies from mRNA vaccines suggests that booster doses can enhance neutralizing antibody response against the Omicron variant and therefore booster doses should be recommended. However, our data suggest that the Omicron variant may be associated with lower COVID-19 vaccine effectiveness against infection even post a third heterologous booster (CoronaVac followed by BNT162b2) and even among previously infected individuals. Our findings have immediate implications for multiples countries that previously used a two-dose regimen of CoronaVac and are of particular importance considering the increasing global need for heterologous vaccine boosters as a relevant future strategy to combat the impact of emerging variants in countries where inactivated vaccines have been the dominant product used.

## Data Availability

All data produced in the present study are available upon reasonable request to the authors.

## Figure Legends

**Table 1: SARS-CoV-2 Vaccinated Cohorts**. Demographic data SARS-CoV-2 Vaccinated Cohort from Dominican Republic (Table1.1) and Yale healthcare workers (HCWs; (Table1.2). Exact counts for each demographic category are displayed in N cell. Percentages of total, where applicable, are displayed in (%) cell. The mean with accompanying standard deviations for each measurement are displayed in Mean and SD cells respectively.

## METHODS

### Ethics statement

This study was approved by the National Bioethics Committee of the Dominican Republic (CONABIOS). The participants received two doses of the inactivated whole-virion vaccine CoronaVac followed by a BNT162b2 booster dose at least four weeks after the second dose of CoronaVac. Republic initiated the COVID-19 vaccination and booster campaigns in February and July 2021, respectively. mRNA vaccine BNT162b2 booster were administrated between July 30 and August 27, 2021. Health care worker (HCW) volunteers from the Yale New Haven Hospital (YNHH) were enrolled and included in this study (IRB Protocol ID 2000028924, approved by the Yale Human Research Protection Program Institutional Review Board. The HCWs volunteers received the mRNA vaccine (100 micrograms mRNA-1273, Moderna or 20 micrograms, BNT162b2, Pfizer-BioNTech) between November 2020 and January 2021. Informed consent was obtained from all enrolled vaccinated. None of the participants experienced serious adverse effects after vaccination.

### Vaccinated volunteers

One hundred and one volunteers from the Dominican Republic and forty HCW participants from the YNHH were followed serially post-vaccination. For the Dominican Republic cohort, plasma samples were collected at baseline (prior to booster, after two doses), 7 and 28 days after the booster (third dose) administration. Plasma from HCWs included on this study were collected 28-days post second vaccination dose. Demographic information was aggregated through a systematic review and was used to construct Table 1. The clinical data were collected using REDCap (v5.19.15 @2021 Vanderbilt University) software. Blood acquisition was performed and recorded by a separate team. Vaccinated clinical information and time points of collection information was not available until after processing and analyzing raw dat. ELISA and neutralizations were performed blinded. Documented history of prior SARS-CoV-2 infection, as confirmed by absence of SARS-CoV-2-specific antibodies, and information of time window post CoronaVac vaccination are available in the Table 1.

### Plasma isolation and storage

Whole blood was collected in heparinized CPT blood vacutainers (BD; # BDAM362780) and kept on gentle agitation until processing. All blood was processed on the day of collection in a single step standardized method. Plasma samples were collected after centrifugation of whole blood at 600 g for 20 min at room temperature (RT) without brake. The undiluted plasma was transferred to 15-ml polypropylene conical tubes, and aliquoted and stored at −80 °C for subsequent shipping and analysis. Plasma samples were sourced from Dominican Republican participants and were shipped to Yale University. The plasma was aliquoted and heat-inactivated at 56°C for 30 min to inactivate complement prior to micro-neutralization.

### SARS-CoV-2 culture

TMPRSS2-VeroE6 kidney epithelial cells were cultured in Dulbecco’s Modified Eagle Medium (DMEM) supplemented with 1% sodium pyruvate (NEAA) and 10% fetal bovine serum (FBS) at 37°C and 5% CO2. The cell line has been tested negative for contamination with mycoplasma. SARS-CoV-2 lineage A (USA-WA1/2020), was obtained from BEI Resources (#NR-52281). Alpha, Beta, Gamma, Delta and Omicron variants were isolated from nasopharyngeal specimens. Alpha, Beta, Gamma, and Delta SARS-CoV-2 samples were sequenced as part of the Yale Genomic Surveillance Initiative’s weekly surveillance program in Connecticut, United States (Kalinich et al., 2020). Omicron (Lineage BA.1) was sequenced by the Connecticut State Department of Public Health (GISAID Accession: EPI_ISL_7313633). The isolates were cultured and resequenced as previously described (Lucas et al., 2021, (Mao et al., 2022). In brief, samples were filtered through a 45μM filter and serially diluted from 1:50 to 1:19,531,250. The dilution was subsequently incubated with TMPRSS2-Vero E6 in a 96 well plate and adsorbed for 1 hour at 37°C. After adsorption, replacement medium was added, and cells were incubated at 37°C for up to 5 days. Supernatants from cell cultures with cytopathic effect (CPE) were collected, frozen, thawed and subjected to RT-qPCR. Fresh cultures were inoculated with the lysates as described above for viral expansion. Nucleic acid was extracted using the ThermoFisher MagMAX viral/pathogen nucleic acid isolation kit and libraries were prepared using the Illumina COVIDSeq Test RUO version. Pooled libraries were sequenced on the Illumina NovaSeq (paired-end 150) by the Yale Center for Genome Analysis. Data was analyzed and consensus genomes were generated using iVar (version 1.3.1). The resequenced genomes were submitted to NCBI (GenBank Accession numbers: ancestral lineage A = MZ468053, Alpha = MZ202178, Beta = MZ468007, Gamma = MZ202306, Delta = MZ468047, Omicron = OL965559). The pelleted virus was then resuspended in PBS and aliquoted for storage at −80°C. Viral titers were measured by standard plaque assay using TMPRSS2-VeroE6. Briefly, 300 μl of serial fold virus dilutions were used to infect Vero E6 cells in MEM supplemented NaHCO3, 4% FBS 0.6% Avicel RC-581. Plaques were resolved at 48h post-infection by fixing in 10% formaldehyde for 1h followed by 0.5% crystal violet in 20% ethanol staining. Plates were rinsed in water to plaques enumeration. All experiments were performed in a biosafety level 3 laboratory with approval from the Yale Environmental Health and Safety office.

### SARS-CoV-2 specific-antibody measurements

ELISAs were performed as previously described (Lucas et al., 2021). Briefly, Triton X-100 and RNase A were added to serum samples at final concentrations of 0.5% and 0.5mg/ml respectively and incubated at room temperature (RT) for 30 minutes before use, to reduce risk from any potential virus in serum. 96-well MaxiSorp plates (Thermo Scientific #442404) were coated with 50 μl/well of recombinant SARS Cov-2 STotal (ACROBiosystems #SPN-C52H9-100ug), or RBD (ACROBiosystems #SPD-C52H3-100ug) at a concentration of 2 μg/ml in PBS and were incubated overnight at 4°C. The coating buffer was removed, and plates were incubated for 1 h at RT with 200 μl of blocking solution (PBS with 0.1% Tween-20, 3% milk powder). Plasma was diluted serially 1:100, 1:200, 1:400 and 1:800 in dilution solution (PBS with 0.1% Tween-20, 1% milk powder) and 100 μl of diluted serum was added for two hours at RT. Human Anti-Spike (SARS-CoV-2 Human Anti-Spike (AM006415) (Active Motif #91351) was serially diluted to generate a standard curve. Plates were washed three times with PBS-T (PBS with 0.1% Tween-20) and 50 μl of HRP anti-Human IgG Antibody (GenScript #A00166, 1:5,000) diluted in dilution solution added to each well. After 1 h of incubation at RT, plates were washed six times with PBS-T. Plates were developed with 100 μl of TMB Substrate Reagent Set (BD Biosciences #555214) and the reaction was stopped after 5 min by the addition of 2 N sulfuric acid. Plates were then read at a wavelength of 450 nm and 570nm.

### Neutralization assay

Sera from vaccinated individuals were heat treated for 30 min at 56°C. Sixfold serially diluted plasma, from 1:10 to 1:2430 were incubated with SARS-CoV-2 variants, for 1 h at 37 °C. The mixture was subsequently incubated with TMPRSS2-VeroE6 in a 12-well plate for 1h, for adsorption. Then, cells were overlayed with MEM supplemented NaHCO3, 4% FBS 0.6% Avicel mixture. Plaques were resolved at 40 h post infection by fixing in 10% formaldehyde for 1 h followed by staining in 0.5% crystal violet. All experiments were performed in parallel with baseline controls sera, in an established viral concentration to generate 60-120 plaques/well.

### Statistical analysis

All analyses of patient samples were conducted using GraphPad Prism 8.4.3 and JMP 15. Multiple group comparisons were analyzed by running parametric (ANOVA) statistical tests. Multiple comparisons were corrected using Tukey’s and Dunnett’s test as indicated in figure legends.

## Acknowledgements

We are very grateful to the study participants who donated specimens for this study. We thank M. Linehan for technical and logistical assistance and D. Mucida for discussions. This work was funded by the Government of the Dominican Republic and supported by the Dominican National Health Cabinet as well as the Ministry of Health. The Dominican Republic team is grateful to Ms Magaly Caram (PROFAMILIA) and Mark Kelly (Laboratorio de Referencia) for their contributions to the set up of the study platform. The study was also supported by the Women’s Health Research at Yale Pilot Project Program (A.I.), Fast Grant from Emergent Ventures at the Mercatus Center (A.I. and N.D.G.), Mathers Foundation, and the Ludwig Family Foundation, the Department of Internal Medicine at the Yale School of Medicine, Yale School of Public Health and the Beatrice Kleinberg Neuwirth Fund. A.I. is an Investigator of the Howard Hughes Medical Institute. C.L. is a Pew Latin American Fellow. V.S.M. is supported by the CAPES-YALE fellowship.

## Author contributions

E.P.T., C.L., V.S.M., M.M, VB, S.V., N.D.G, and A.I. conceived the study. S.O., A.K., and IY designed and implemented HCW cohort study. V.B., L.C., E.C., A.J., M.S., I.Yand P.L collected and processed plasma samples. C.L. and V.M.S. performed SARS-CoV-2 specific antibody ELISAs and the neutralization assays. VB, LC, M.C. and I.Y., collected epidemiological and clinical data. C.B.F.V., M.I.B., K.B., C.P., R.D., E.G., A.M., J.R., surveilled, detected and performed virus sequencing. C.L and V.S.M., isolated SARS-CoV-2 variants. VB, I.Y., and M.C., assisted volunteers’ identification and enrolment. C.L., and V.M.S. analyzed the data. EPT and MM designed, managed and control the quality of epidemiological data collected at REDCap database. C.L., E.P.T., and A.I. drafted the manuscript. All authors reviewed and approved the manuscript. E.P.T and A.I. secured funds and supervised the project.

